# Perceived vs. actual navigation ability: Differences between autistic and typically developing children

**DOI:** 10.64898/2026.04.09.26350542

**Authors:** Daniel J. McKeown, Oscar S. Cruzado, Giorgio Colombo, Douglas J. Angus, Victor R. Schinazi

**Affiliations:** School of Psychology, Faculty of Society and Design, Bond University, Gold Coast, Queensland, 4229, Australia; Future Health Technologies, Singapore-ETH Centre, Campus for Research Excellence and Technological Enterprise (CREATE), Singapore

**Keywords:** Spatial cognition, Neurodevelopment, Sense of direction, Self-perception

## Abstract

**Purpose:** Navigational ability develops throughout childhood alongside the maturation of brain regions supporting egocentric and allocentric processing. In Autism Spectrum Disorder (ASD), atypical hippocampal development may impact flexible spatial memory; however, findings on navigational ability in autistic children remain inconsistent. This study aimed to compare both objective and perceived navigation ability in children with ASD and typically developing (TD) peers.

**Method:** Twenty-six children with high-functioning ASD and twenty-five age- and gender-matched TD children (M_age = 12.04 years, SD = 1.64) completed a battery of navigational tasks from the Spatial Performance Assessment for Cognitive Evaluation (SPACE), including Path Integration, Egocentric Pointing, Mapping, Associative Memory, and Perspective Taking. Perceived navigation ability was assessed using the Santa Barbara Sense of Direction (SBSOD) scale.

**Results:** No significant group differences were observed across any objective navigation tasks. However, children with ASD reported significantly lower perceived navigation ability compared to TD peers.

**Conclusion:** These findings suggest a dissociation between perceived and actual navigational ability in ASD. By early adolescence, objective navigation performance appears intact, potentially reflecting sufficient maturation of underlying neural systems or the presence of compensatory mechanisms. The results underscore the importance of incorporating objective, task-based measures when assessing cognitive abilities in autistic populations.

## Perceived vs. actual navigation ability: Differences between autistic and typically developing children

The ability to navigate accurately through an environment is essential for daily functioning and survival (Lee & Spelke, 2010; Newcombe, 2019). Whether determining the shortest route to work or adjusting to an unexpected detour, effective navigation relies on cognitive processes that integrate sensory information acquired from a first-person perspective into representations of one’s position and orientation in space. Many of the cognitive processes that support navigation in humans are not fully developed at birth but emerge progressively throughout childhood and adolescence (Bullens et al., 2010; Nardini et al., 2006; Ribordy et al., 2013). This developmental trajectory reflects the gradual maturation of neural systems (e.g., hippocampus and prefrontal cortex), which underpin spatial memory, route planning, and flexible adaptation to environmental changes. Understanding how these abilities develop provides important insights into both typical cognitive growth and is critical for determining whether, and in what ways, navigation abilities differ between neurotypical individuals and those with autism spectrum disorder (ASD).

During navigation, individuals may encode spatial information using egocentric and allocentric reference frames (Hartley et al., 2003; Lee & Spelke, 2010). In an egocentric reference frame, spatial information is encoded relative to the individual’s own position and orientation (e.g., the church is to my left) and is often associated with response-based strategies such as learning a sequence of actions or movements to reach a goal (Burgess, 2006; Gramann et al., 2010; Iglói et al., 2009). In an allocentric reference frame, spatial information is encoded relative to external landmarks, independent of the individual’s position (e.g., the church is west of the mall), and is associated with place-based strategies that support the formation of map-like representations of space (Burgess, 2006; Gramann et al., 2010; Marchette et al., 2014; O’Keefe, 1991).

Findings from animal and human studies highlight the use of both strategies (Ekstrom, 2018; Colombo et al., 2017; Ekstrom et al., 2014; Epstein et al., 2017; Klatzky, 1998; Montello, 2005). In unfamiliar environments, response-based strategies are often used first, with place-based strategies becoming more efficient once the environment is familiar (Brake & Lacasse, 2018; Hartley et al., 2003). Here, classic maze experiments have found that animals trained to follow a fixed sequence of turns from a consistent starting point can reach the goal even when distal cues are removed, consistent with response-based navigation (Morris, 1984; Morris, 1981; Tolman, 1948). Similar patterns are observed in human virtual maze studies, where participants sometimes adopt habitual turn sequences even when shortcuts are available, particularly under stress or time pressure (Babayan et al., 2017; Bohbot et al., 2007; Brown et al., 2012; Gillner & Mallot, 1998; Iaria et al., 2003; Konishi & Bohbot, 2013). Other findings show that animals and humans can take novel shortcuts and infer the location of landmarks they have not directly visited, indicating the use of a place-based strategy supported by a cognitive map (Alvernhe et al., 2008; Baumann et al., 2012; Erdem & Hasselmo, 2012; Javadi et al., 2019; Pazzaglia et al., 2018).

In humans, children tend to rely more on response-based strategies and perform worse than adults on wayfinding tasks, particularly when routes are complex or landmarks are absent or unlabelled (Jansen-Osmann & Wiedenbauer, 2004; Lingwood et al., 2015; Nys et al., 2014; Sneider et al., 2015). Place-based strategies are also less developed in children, who often need to move physically through an environment to perform well (Ebersbach et al., 2011; Newcombe & Frick, 2010; Pearson et al., 2013). While flexibility in switching between response and place strategies improves during adolescence, the development of these strategies does not occur concurrently and is in conflict during the early stages of life (Brucato et al., 2022; Sneider et al., 2015). Place-based learning emerges from age two and progressively strengthens throughout the early years of life (Balcomb et al., 2011; Ribordy et al., 2013). This development occurs concurrently with structural and functional maturation of medial temporal structures, particularly the hippocampus (Newcombe et al., 1998; Reinhardt et al., 2020). At the neuronal level, hippocampal place cells encode specific locations within the environment and integrate these into a coherent spatial framework that supports spatial inference and flexible navigation (Gillner & Mallot, 1998; Newcombe, 2019). However, the trajectory for hippocampal development is not uniform across all individuals (Reinhardt et al., 2020). In cases of atypical maturation, neurodevelopmental conditions can lead to significant alterations in cognitive function and spatial learning.

ASD is characterised by atypical cognitive abilities that result in changes in social skills, speech and language, and the development of repetitive and restricted activities (e.g., motor sterotypies, preoccupation with unusual objects and routines; Hirota & King, 2023). Together, these characteristics can negatively affect every day functioning, including the ability to navigate and adapt to complex environments (Smith, 2015). Although restricted and repetitive behaviours associated with ASD might be expected to influence spatial and navigational skills, findings are mixed, with studies reporting impairment, improvement, or no differences relative to Typically Developing (TD) individuals. For example, researchers have found that High-Functioning ASD (HFASD) children have impaired performance when required to locate objects in a three-dimensional simulated environment (Lind et al., 2014), perform large-scale search behaviour (Pellicano et al., 2011) and active exploration (Fornasari et al., 2013; Pierce & Courchesne, 2001) in a purpose-built environment. Conversely, HFASD children have also demonstrated comparable or superior performance to TD children when performing spatial perspective taking (Presley, 2021; Hobson, 1984; Kim et al., 2013; Reed & Peterson, 1990), maintaining spatial relationships of objects in a virtual environment (Edgin & Pennington, 2005), judging distances to objects from a first-person perspective (Giovannini et al., 2009), and route learning in a virtual environment (Arnold et al., 2014; Caron et al., 2004; Chrastil et al., 2015; Edgin & Pennington, 2005; Fornasari et al., 2013; Laidi et al., 2023; Noel et al., 2020; Shrager et al., 2008).

Spatial and navigation performance may be proportional to the degree of preserved cognitive ability (i.e., executive functioning, memory, motor control; Bangerter et al., 2017; Berenguer et al., 2018; Hemmers et al., 2022; Ming et al., 2007) and the integrity of brain regions that are critical for navigation (He et al., 2025; Holiga et al., 2019). In individuals with ASD, Magnetic Resonance Imaging (MRI) studies have consistently demonstrated atypical structural development in temporal, frontal, cerebellar, and hippocampal brain regions (Chaddad et al., 2017; He et al., 2025; Holiga et al., 2019; Laidi et al., 2017), as well as functional dysconnectivity between these regions (Cristian Gonzalez-Rodriguez, 2025; Igelström et al., 2017; Khan et al., 2015; Stoodley et al., 2017). Because hippocampal maturation occurs concurrently with the emergence of early ASD symptoms, the hippocampus has been mechanistically linked to the pathophysiology of ASD (Banker et al., 2021). Hippocampal volume is reduced in ASD compared to TD individuals, especially in CA1 regions important for navigation (Banker et al., 2021). Furthermore, the severity of reduction is correlated with the severity of ASD traits and cognitive deficit (Arutiunian et al., 2023; Eilam-Stock et al., 2016). Notably, if hippocampal growth can be maintained throughout maturation, the severity of ASD traits can be reduced (Reinhardt et al., 2020).

The functional characteristics of the hippocampus also differ in ASD, with reductions in synaptic complexity contributing to increased cognitive deficits. Here, reductions in synaptic complexity have been associated with impaired memory retrieval and encoding of allocentric environmental information (Gaigg et al., 2015; Patil et al., 2024; Urbain et al., 2015). Additionally, supportive structures for hippocampal activity, such as the cerebellum, are altered in individuals with ASD (Kelly et al., 2020; Stoodley et al., 2017). The cerebellum aids in optimising hippocampal representations of an environment (Rondi-Reig et al., 2022). However, cerebrocerebellar circuits in ASD appear to be intact in some cases (Laidi et al., 2023; Traut et al., 2018). For example, during navigation of a Starmaze task, adults with ASD showed performance comparable to TD adults, which coincided with similar cerebellar Crus I volume and functional connectivity (Laidi et al., 2023). In parallel, response-based navigation strategies mediated by the caudate nucleus appear to remain relatively intact in ASD (Caron et al., 2004; Lind et al., 2013). This has been demonstrated in corridor-based and maze-like navigation tasks, where individuals with HFASD perform similarly to, or in some cases better than, TD individuals. In these contexts, children with ASD show preserved spatial working memory, route learning, and map learning, suggesting that non-hippocampal navigation systems can support successful performance (Caron et al., 2004; Edgin & Pennington, 2005; Laidi et al., 2023).

Many commonly used navigation tasks lack sensitivity to the multiple cognitive components that jointly support navigation. Such assessments are also often constrained by strict environmental boundaries (e.g., maze walls) and can be difficult to administer in children with ASD (Laidi et al., 2023). These shortcomings highlight the need for accessible assessment tools that can probe multiple, dissociable facets of navigation within a single framework, allowing deficits and compensatory mechanisms to be examined across tasks rather than inferred from a single performance outcome. The Spatial Performance Assessment for Cognitive Evaluation (SPACE) is a serious iPad-based tool designed to assess multiple facets of spatial navigation (Colombo, Minta, Grübel, et al., 2024). In SPACE, participants complete five tasks (i.e., path integration, egocentric pointing, mapping, associative memory, and perspective taking) that differentially rely on egocentric and allocentric strategies. SPACE has demonstrated strong reliability (Tian et al., 2025) and validity as a navigation assessment tool and has been validated across both healthy and clinical populations. Performance in SPACE has been shown to discriminate between different levels of cognitive impairment (Colombo et al., 2026) and to relate to hippocampal structure, with better performance associated with larger hippocampal volume (Minta et al., 2026). In addition, extensive normative data are available across adulthood (approximately 20 to >60 years), accounting for both age and gender effects (Colombo et al., 2024). Together, these properties position SPACE as a well-suited tool for characterising navigation abilities and strategy use in younger individuals with HFASD.

In the current study, we used SPACE to compare the navigational ability of children with HFASD to TD children. We hypothesise that children with ASD will demonstrate strengths in tasks involving egocentric and route learning, while expecting impairments in tasks requiring allocentric knowledge and the adoption of a different perspective. As such, we anticipate superior or intact performance in the path integration and egocentric pointing tasks, while expecting diminished abilities in the mapping and perspective taking tasks.

## Methods

### Participants

We recruited 26 children diagnosed with ASD (M_age_ = 12.04 ± 1.64, 5 female). This group of participants were recruited from a specialist school in Brisbane, Queensland, Australia. Participants had received a formal diagnosis of ASD according to the DSM-5 criteria and were considered high-functioning (DSM severity level: Level 1; IQ > 70). The DSM categorises the severity of ASD across three levels, with Level 1 indicating that the individual “requires support” to manage deficits in social communications, switching between activities, and restrictive/repetitive behaviours (American Psychiatric Association, 2013). Given the high prevalence of attention-deficit/hyperactivity disorder (ADHD) in individuals with ASD (Eaton et al., 2023), we deliberately included participants with ADHD to ensure a more inclusive and representative sample of the autistic population. The comparison group consisted of 25 TD children (M_age_ = 12.12 ± 1.59, 4 female), matched on chronological age and gender. All participants were recruited from secondary schools in Brisbane, Australia. All participants were fluent in English and reported no history of cognitive or psychiatric disorders outside of the specified inclusion criteria. Participants gave written and verbal consent before partaking in the study. Ethical approval for the study was obtained from the Bond University Human Research Ethics Committee (ethics approval number: VS0306) and was performed in accordance with the ethical standards as laid down in the 1964 Declaration of Helsinki. Informed consent was obtained from both parents and children prior to testing. All participants received a $50 gift card upon completion of the study.

### Sociodemographic and Health Questionnaires

Participants completed a comprehensive questionnaire designed to gather sociodemographic and health-related information. This included age, educational background, handedness, prior experience using tablets, and any formal navigation training. The questionnaire also collected general health information, such as visual status (e.g., impairments or corrections), chronic medical conditions, and history of traumatic brain injury. Additionally, participants reported on their daily sleep duration and weekly physical activity levels, including time spent walking and engaging in vigorous exercise. Sociodemographic and health characteristics are reported in Table 1 in Supplementary Material 1.

### The Santa Barbara Sense of Direction (SBSOD)

The SBSOD is a 15-item self-report measure designed to assess an individual’s perceived navigation ability (Hegarty et al., 2002). Participants rate their agreement with statements such as “I am very good at giving directions”, “I enjoy reading maps”, and “It is not important to me to know where I am” using a 7-point Likert scale. Final scores were computed by averaging item responses, with higher scores indicating a stronger self-perceived sense of direction. The SBSOD scale has demonstrated high internal consistency (α = .87 - .88; Davies et al., 2017) and excellent test-retest reliability (r = .91; Hegarty et al., 2002).

### Spatial Performance Assessment for Cognitive Evaluation (SPACE)

SPACE (Colombo et al., 2026; Colombo, Minta, Thrash, et al., 2024; Minta et al., 2026; Tian et al., 2025) is an iPad-based serious game developed to evaluate individual differences in spatial navigation. In SPACE, participants assume the role of an astronaut tasked with exploring a new planet. The game includes a training phase followed by five spatial tasks: path integration, egocentric pointing, mapping, associative memory, and perspective taking (Figure 1).

**Figure 1.**
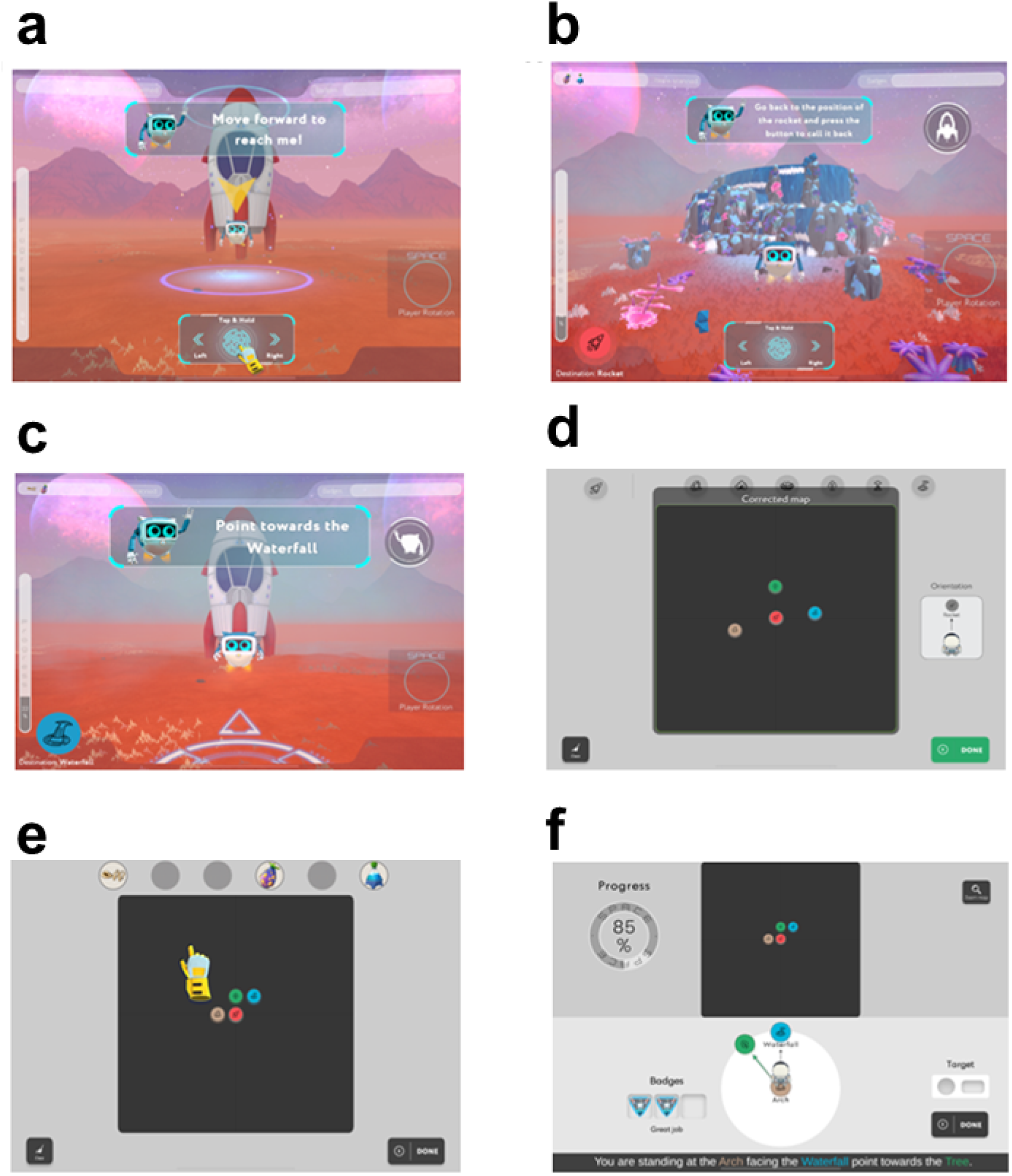
Each task of the Spatial Performance Assessment for Cognitive Evaluation (SPACE). During the training phase (a), participants learn basic controls to translate through the environment. In Path Integration (b), participants are required to return to the rocket’s location unaided after visiting two landmarks in the environment. Egocentric Pointing (c) requires participants to estimate the locations of landmarks in the environment in relation to another landmark. Participants are then required to build a cognitive map during the Mapping task (d). In the Associative Memory tasks (e), participants are required to pair up items they picked up during Path Integration at their respective landmarks. Finally, participants use a map of the environment to estimate the locations of landmarks relative to other landmarks in a top-down perspective during the Perspective Taking task (f).

The training phase serves to familiarise participants with the SPACE interface and controls, and to prepare them for subsequent SPACE tasks, while concurrently assessing basic motor and visuospatial functions (Figure 1A). During this phase, participants learn to rotate and translate in the virtual environment by following a robot to various destinations. Upon successful completion of the training phase, participants proceed to the path integration task. In the path integration task, participants complete a series of triangle-completion tasks designed to assess their ability to monitor changes in their position and orientation as they move through the environment. Each trial begins at a visible starting point (e.g., the rocket), which takes off immediately at the onset of the trial and is therefore no longer visible during navigation. Participants are then guided by a robot from the starting location to two successive landmarks. At each landmark, the robot scans a different element that will be recalled in a later task. After reaching the second landmark, participants are instructed to return to the starting point (e.g., the rocket) using the most direct route. This return leg is performed without guidance and in the absence of the starting cue, requiring reliance on internal spatial updating processes. In path integration, the spatial configuration of the landmarks remains consistent across seven trials (Figure 1b).

The egocentric pointing task assesses participants’ memory for the spatial layout encountered during the path integration task from a first-person perspective. Over a series of trials, participants are positioned in front of different landmarks or the starting location (e.g., the rocket) and asked to point in the direction of previously visited landmarks (Figure 1c). Each response requires participants to recall the locations of the landmarks and determine their direction in the absence of visual cues. The mapping task requires participants to reconstruct the spatial layout of the environment from a top-down perspective. Participants are required to drag and drop the icons of the landmarks they previously navigated to in the path integration task onto an empty canvas with the goal of building a map of the virtual environment (Figure 1d). The associative memory task assesses participants’ ability to match items (e.g., water) previously scanned by the robot during the path integration task with their corresponding landmarks (e.g., waterfall, tree; Figure 1e). Perspective taking task tests an individual’s ability to judge relationships between landmarks while alternating perspectives. Participants are aided with a top-down map of the environment and are asked to maintain the perspective of standing at a landmark while facing another landmark. Participants are then required to estimate the position of a third landmark (e.g., “imagine you are standing at the tree facing the waterfall. Point towards the arch”; Figure 1f).

For the training phase, performance is quantified as the overall time to complete the various phases of training in seconds. Better performance is indicated by a shorter completion time. For path integration, performance is quantified as the distance between the participant’s final position and the target position, in game units. Better performance is indicated by smaller distance error. For the egocentric pointing task, performance is quantified as the absolute angular deviation in degrees for the participant’s judged position of a landmark and the actual position of the landmark. Better performance is indicated by smaller angular errors. For the mapping task, performance is quantified using bidimensional regression (Friedman & Kohler, 2003; Tobler, 1994) with R^2^ values closer to 1 indicating better mapping accuracy. In the associative memory task, performance is quantified by the percentage of correct landmark-object pairings. Better performance is indicated by larger percentage values. For the perspective taking task, performance is quantified by the absolute angular deviation in degrees of the participant’s judged position of a landmark and the actual position of the landmark. Better performance is indicated by smaller angular errors. Performance measures were averaged across all trials for each task.

### Experimental Procedure

Prior to the experiment, parents were contacted via email and/or telephone to explain the study’s purpose, procedures, and ethical considerations. An explanatory statement detailing participants’ rights and study information was provided. Informed consent was then obtained from both the parent and the participant. Participants in the ASD and TD groups were assessed in a designated testing room at their school. All participants completed the sociodemographic and health questionnaire and the SBSOD prior to completing SPACE. In SPACE, participants completed the training phase followed by the path integration, pointing, mapping, associative memory and perspective tasks. The task order was not randomised to preserve the game’s story, which requires participants to complete the path integration task to learn the positions of the landmarks in the virtual environment. Each task was introduced with a verbal explanation, followed by a brief instructional video displayed on the iPad. Upon completing the session, participants were debriefed and invited to ask questions or share feedback with the researcher.

### Statistical Analysis

All statistical analyses were performed in Jamovi (v 2.6.13) and the MASS package for robust regressions in RStudio. Before assessing differences between the ASD and TD groups, Pearson correlations were computed to examine the relationships among age, SBSOD score, and performance during the SPACE task.

#### Objective navigation performance

For each SPACE task, task-specific performance measures were analysed using separate linear regression models. Age (continuous) and Condition (TD vs ASD) were entered simultaneously as predictors to assess developmental effects and group differences in navigation performance. Model assumptions were evaluated by inspecting residual distributions and formally tested using the Shapiro-Wilk test for normality, and other diagnostic parameters were measured prior to interpreting model outputs (i.e., Cook’s distance, Durban-Watson Test for Autocorrelation, Collinearity). If normality or diagnostics were violated, estimates were corrected using robust linear models. Given that all participants achieved perfect performance on the associative memory task (100% accuracy), this task was excluded from further analyses. Additionally, observed performance distributions were examined relative to these reference values to assess whether the current sample performed within the expected normative range.

#### Subjective navigation ability

Subjective spatial orientation ability was assessed using the SBSOD scale. Differences between groups in overall SBSOD scores and individual SBSOD items were assessed using separate linear regression models. Age (continuous) and Condition (TD vs ASD) were entered simultaneously as predictors to assess developmental effects and group differences in subjective navigational ability. Similar to the analysis of objective navigation performance, estimates from robust linear models were reported if normality or diagnostics were violated.

## Results

### Correlation between SPACE performance and Subjective Navigational Ability

Pearson correlations were performed to assess the relationships among participant age, SBSOD score, and SPACE performance measures (Figure 1 in Supplementary Material 1). Significant correlations were found between Path Integration Distance Error and Egocentric Pointing Error (*r* = .31, *p* = .03), Path Integration Distance Error and Perspective Taking Error (*r* = .31, *p* = .03), Egocentric Pointing Error and Mapping (*r* = -.64, *p* < .001), and Egocentric Pointing Error and Perspective Taking Error (*r* = .33, *p* = .02). Notably, there was a significant negative relationship between age and Path Integration distance error (*r* = -.28, *p* = .04), and Age and Perspective Taking error (*r* = -.32, *p* = .02), such that older participants had smaller errors during the Path Integration and Perspective Taking tasks than younger participants. There were no significant correlations between Age and the other SPACE tasks or SBSOD score.

### SPACE Performance

Descriptive statistics are reported in Table 2 in Supplementary Material 1, and SPACE performance measures are shown in Figure 2. For Path Integration Distance Error, the overall model was not significant, *F*(2, 48) = 2.09, *p* = .134, *R*² = .04. Age was a significant predictor, β = -8.8, SE = 4.31, *t* = -2.04, *p* = .047, indicating that older children performed better in Path Integration. Condition was not a significant predictor, β = .96, SE = 13.65, *t* = .07, *p* = .944. Because the model residuals violated the assumption of normality, *p* = .004, a robust linear model using Huber’s M-estimator was also conducted. The effect of Age remained consistent, β = -7.50, SE = 3.72, robust *t* = -2.01.

**Figure 2.**
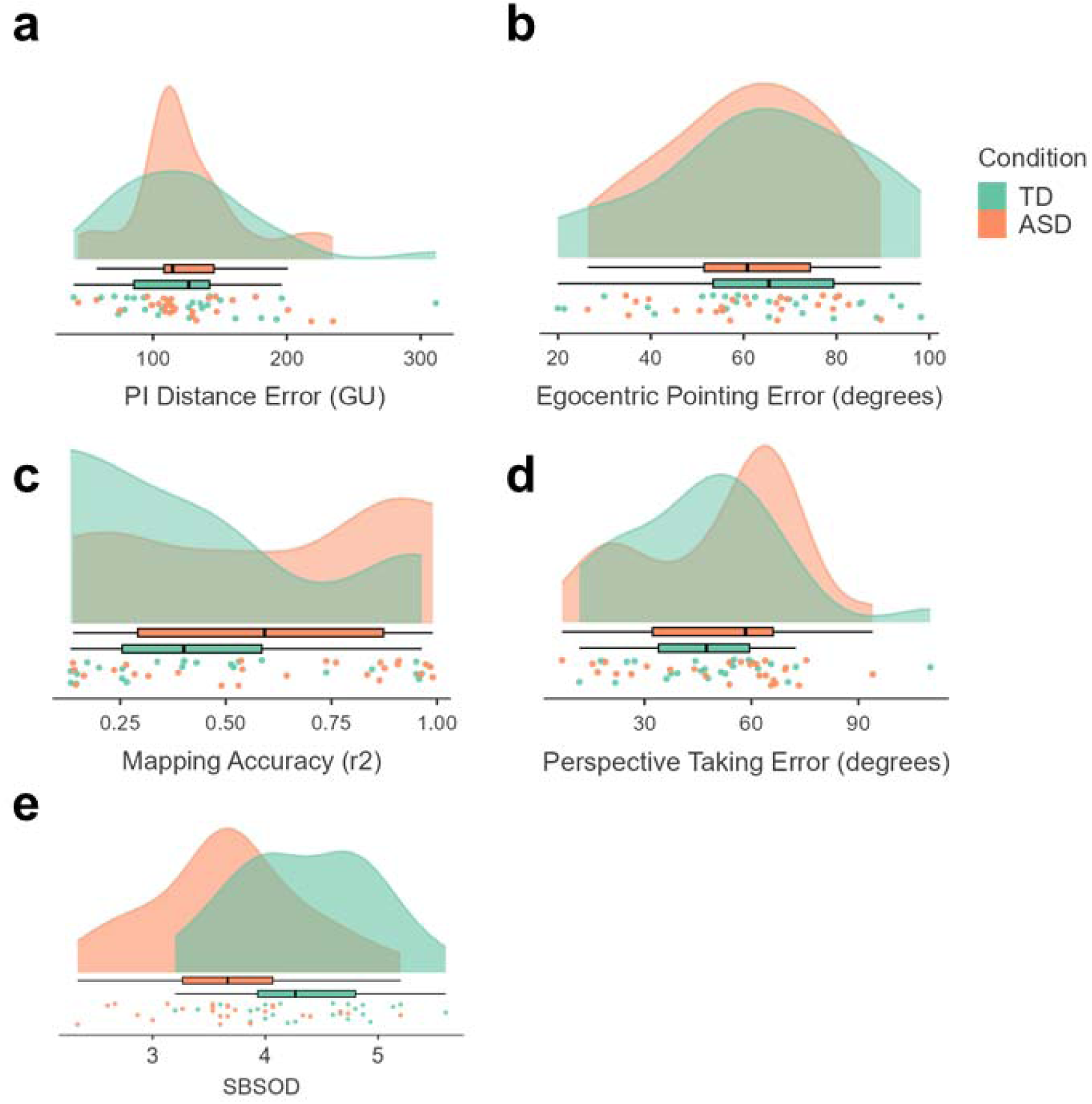
Performance measures for each task of the Spatial Performance Assessment for Cognitive Evaluation (SPACE) for autism spectrum disorder (ASD) and typically developed (TD) children. There were no differences between groups for Path Integration (PI; a), Egocentric Pointing (EP; b), Mapping (c), or Perspective Taking (PT; d) tasks. GU = game units. Santa Barbara Sense of Direction (SBSOD) scale score for autism spectrum disorder (ASD) and typically developed (TD) children (e). Box plot represents median and quartiles.

For Egocentric Pointing Error, the overall model was not significant, *F*(2, 48) = 0.14, *p* = .863, *R*² < .01. Age and Condition were not significant predictors, Age: β = -.06, SE = 1.73, *t* = -.04, *p* = .969; Condition: β = -2.98, SE = 5.50, *t* = -.54, *p* = .591. For Mapping, the overall model was also not significant, *F*(2, 48) = 1.45, *p* = .243, *R²* < .01. Age and Condition were not significant predictors, Age: β = -.015, SE = .026, *t* = -.571, *p* = .571; Condition: β = .134, SE = .08, *t* = 1.59, *p* = .117.

For Perspective Taking Error, the overall model was not significant, *F*(2, 47) = 2.94, *p* = .062, *R*² = .07. Age was a significant predictor, β = -4.39, SE = 1.87, *t* = -2.35, *p* = .023, indicating that older children performed better in Perspective Taking. Condition was not a significant predictor, β = 3.23, SE = 5.98, *t* = .54, *p* = .592. Because the model residuals violated the assumption of normality p = .04, a robust linear model using Huber’s M-estimator was also conducted. The effect of Age remained consistent, β = -4.82, SE = 1.70, robust *t* = -2.83.

### Subjective Navigational Ability

Descriptive statistics are reported in Table 3 in Supplementary Material 1, and SBSOD total scores are shown in Figure 2e. For the SBSOD total score, the overall model was significant, *F*(2, 47) = 9.32, p < .001, R^2^ = .253. Condition was a significant predictor, β = -.71, SE = .18, *t* = -3.95, *p* < .001, but Age was not, β = -.11, SE = .06, *t* = -1.95, *p* = .057, indicating that TD children reported higher subjective navigational ability than ASD children. Regarding the individual items of the SBSOD, the linear regression model only explained a significant amount of variance for Item 1 “*I am very good at giving directions”*, *F*(2, 47) = 13.5, p < .001, R^2^ = .34 and Item 5 *“I tend to think of my environment in terms of cardinal directions (N, S, E, W)”*, *F*(2, 47) = 9.51, p <.001, R^2^ = .26. For Item 1, Condition was a significant predictor, β = -1.62, SE = .32, *t* = -5.07, *p* <.001, but not Age, β = -.14, SE = .10, *t* = -1.37, *p* = .177. For Item 5, Condition and Age were significant predictors, Condition: β = - 1.93, SE = .49, *t* = -3.92, *p* < .001; Age: β = -.33, SE = .16, *t* = -2.11, *p* = .04. For these items, TD children scored their subjective navigational ability higher than ASD children specifically for these items of the SBSOD.

## Discussion

In the current study, we aimed to identify how children with ASD perform on a suite of tasks designed to detect deficits in navigational ability, compared with TD children. Twenty-six otherwise healthy children with ASD and 25 TD children completed the Path Integration, Egocentric Pointing, Mapping, and Perspective-Taking tasks in SPACE. We hypothesised that children with ASD would perform worse than TD children on tasks emphasising allocentric navigation. Surprisingly, children with ASD performed just as well as children with TD across all SPACE tasks. However, children with ASD rated their self-perceived navigation ability to be worse than that of TD children on the SBSOD scale.

### Actual navigational ability is maintained in ASD

Both ASD and TD children performed comparably well on the Path Integration, Egocentric Pointing, Mapping, and Perspective-Taking tasks. This supports the notion that navigational ability in ASD is not impaired compared to TD children (Giovannini et al., 2009; Laidi et al., 2023; Noel et al., 2020; Warreyn et al., 2005). Navigation involves solving spatial problems by using allocentric and egocentric strategies. Because of this, navigation reflects a continuum where individuals can shift between, or simultaneously draw upon, allocentric and egocentric representations to successfully complete spatial problems. Considering the extensive structural and functional connections among brain regions involved (Baumann & Mattingley, 2021; Brown et al., 2012), different neural systems involved in navigation can be recruited interchangeably depending on task demands (Burgess, 2006). This is supported by studies showing that performance in path integration (Arnold et al., 2014; Caron et al., 2004; Chrastil et al., 2015; Edgin & Pennington, 2005; Fornasari et al., 2013; Laidi et al., 2023, 2023; Noel et al., 2020; Shrager et al., 2008), mapping (Bohbot et al., 1998; Caron et al., 2004), and perspective taking (Amber Presley, 2021; Hobson, 1984; Kim et al., 2013; Reed & Peterson, 1990) task can still be performed accurately in the presence of impaired structure and function of the hippocampus.

In SPACE, the Path Integration task requires participants to engage egocentric and allocentric strategies to update self-motion relative to the origin point, and to learn the positions of the various landmarks for subsequent tasks. This process is primarily supported by medial temporal regions, including the hippocampus and entorhinal cortex, and, to a degree, by parietal and vestibular networks (Arnold et al., 2014; Epstein et al., 2017; Segen et al., 2022). Egocentric pointing requires participants to retrieve and update self-referenced spatial information to estimate the direction of unseen landmarks from a first-person perspective, engaging posterior parietal regions for egocentric updating and retrosplenial regions to coordinate the transformation. The Mapping task emphasises an individual’s ability to integrate landmark information from previous tasks and to form an allocentric representation of the environment, the success of which is strongly linked to hippocampal and retrosplenial activity (Wolbers & Buchel, 2005). However, the mapping task can be completed using egocentric strategies by reconstructing the route between landmarks to form a map, drawing upon basal ganglia function (Iaria et al., 2003; Schinazi et al., 2013). Lastly, the Perspective Taking task requires participants to adopt allocentric and egocentric strategies to perform transformations between landmarks and flexibly manipulate their spatial representations to form alternative perspectives, engaging parietal and medial temporal neural networks, as well as prefrontal cortices (Lambrey et al., 2012; Schinazi et al., 2013). Considering this, the lack of differences in navigational ability between ASD and TD participants observed in the current study may reflect compensatory effects of other brain networks masking navigational impairment, even if the hippocampus is atypical in ASD.

### Age-dependent improvements in navigational ability

Younger age was associated with worse performance on the Path Integration and Perspective Taking tasks, but not the Egocentric Pointing or Mapping tasks. Interestingly, these effects emerged within a relatively restricted age/developmental window, suggesting that measurable improvements in navigational ability continue to develop even across late childhood. Furthermore, this pattern indicates selective improvement in SPACE tasks that require individuals to dynamically update spatial representations and perspectives in later childhood.

The absence of age effects during egocentric pointing is unsurprising, considering that this task primarily relies on egocentric strategies that may develop earlier in childhood (Jansen-Osmann & Wiedenbauer, 2004; Lingwood et al., 2015; Nys et al., 2014; Sneider et al., 2015). However, the interpretation of the absence of age effects during Mapping is not as clear. As noted previously, it is possible to complete mapping-like tasks using other strategies, such as implementing response-based strategies that rely less on flexible egocentric framing (Burgess, 2006). Here, TD and ASD participants have not reached a developmental threshold at which the implementation of allocentric strategies results in improved mapping beyond that of response-based strategies. Considering this, the absence of differences in mapping performance between groups should not be interpreted as evidence of maturation in mapping ability across groups but rather may reflect the variability in strategies used to complete the Mapping task (Brucato et al., 2022; Nazareth et al., 2018).

Although path integration is typically characterised as an egocentric process, it also requires the individual to maintain landmark associations over time as they move through an environment (Epstein et al., 2017). Improvements across this small age range may therefore reflect maturation in multiple processes involved in dynamic spatial updating that contribute to the successful integration of multiple sensory inputs and to working memory, which depends on parietal and medial temporal lobe interactions (Balcomb et al., 2011; Brucato et al., 2022; Sneider et al., 2015). Similarly, the age effect observed in Perspective Taking is consistent with previous evidence that shows progressive improvement in the ability to adopt and transform alternative viewpoints throughout childhood (Bullens et al., 2010; Cardillo et al., 2020; Ebersbach et al., 2011; Frick et al., 2014; Hollarek & Lee, 2022; Nardini et al., 2006; Newcombe, 2019). Perspective-taking requires an individual to imagine different orientations within the environment and to infer the positional relationships among landmarks (Frick et al., 2014). Although children can adopt alternative perspectives from early childhood when a stable reference point is provided, performance declines when transformations must be generated flexibly or vary trial to trial (Hegarty & Waller, 2004; Hollarek & Lee, 2022; Huttenlocher & Presson, 1979).

To further support this, normative data for SPACE performance have recently become available (Colombo, Minta, Thrash, et al., 2024). Here, both the TD and ASD participants performed within expected adult error ranges for 21 to 29-year-olds in Path Integration (75-90^th^ percentile), Egocentric Pointing (50^th^ percentile), and Mapping (50^th^ percentile), whereas Perspective-Taking performance was weaker in comparison and corresponded to the equivalent of the 25-50^th^ percentile of the 60+ age group. These comparisons further support our claim that the ability to use flexible allocentric transformations continues to develop throughout late childhood.

### Self-perceived navigational ability is impaired in ASD

While there were no differences in performance between ASD and TD children during SPACE, self-reported ability to navigate was worse for ASD children compared to TD children. Furthermore, self-reported navigational ability was not correlated with performance on the SPACE tasks. This dissociation suggests that the reduced SBSOD scores observed in the ASD group likely reflect differences in self-confidence in their navigational ability rather than objective differences in navigational competence.

Children with ASD may underreport their general navigational abilities. The SBSOD scale assesses broad, trait-like perceptions of everyday navigation rather than performance in specific navigation domains (Hegarty et al., 2002). Spatial anxiety has previously been associated with self-reported navigational ability and greater reliance on route-based strategies (Farran et al., 2022; Mueller et al., 2009; Pazzaglia et al., 2018). Anxiety disorders are also more prevalent in ASD compared to TD populations (van Steensel et al., 2011), which increases the probability that heightened anxiety in the ASD participants could have contributed to the lower SBSOD scores (Varshney et al., 2024). However, as we did not observe an association between SBSOD scores and performance during any of the SPACE tasks, reduced self-perceived navigational ability did not impair actual navigational performance. This is important as our findings demonstrate that these beliefs don’t translate into measurable performance decrements when individuals are required to navigate in a controlled environment.

The individual SBSOD item analysis offers insight into the nature of this reduced self-perceived navigational ability seen in ASD individuals. Items 1 and 5 were primarily responsible for the group differences in SBSOD scores. Item 1 (“*I’m very good at giving directions”*) may reflect broader confidence when needing to make momentary adaptations in navigation, whereas Item 5 (*“I tend to think of my environment in cardinal directions”*) may indicate confidence with map-based representations of an environment (Davies et al., 2017). ASD individuals tend to prefer familiar, habitual, or route-based strategies, especially under stress or heightened anxiety (Smith, 2015). This may explain the presence of lower scores in the ASD group on these items, even though they are still capable of effectively navigating using alternative strategies. However, this interpretation is cautious, as reductions in SBSOD scores may also reflect differences in personality traits, which can confound responses beyond navigational ability (Condon et al., 2015). These findings highlight a potential methodological issue of using self-report measures to categorise navigational ability in ASD. Here, the SBSOD survey-based tool has reported spatial ability differences in ASD populations, whereas a performance-based tool such as SPACE shows that navigational ability is indeed intact.

### Considerations

The ASD cohort included a large proportion of participants who also had a co-occurring ADHD diagnosis. Given that ADHD and ASD diagnoses commonly overlap, it was not possible to separate our ADHD participants into those with and without an ADHD diagnosis and maintain statistical power to isolate ADHD-related effects. Future research should aim to do this, as attentional control, executive functioning, and impulsivity associated with ADHD may impact navigational ability (Bora & Pantelis, 2016). The restricted age range in our sample may have also limited the ability to detect more pronounced differences in navigation. Although age effects were observed for Path Integration and Perspective-Taking, performance in these tasks fell within expected normative ranges (Colombo, Minta, Thrash, et al., 2024). Future research could manipulate the difficulty of the tasks in SPACE to better characterise developmental differences through late childhood. While we suggest that some features of navigational impairment in our cohort may be due to underdevelopment of hippocampal function, we are unable to definitively demonstrate that the structure and function of the hippocampus are similar between our TD and ASD cohorts. Considering this, future research should incorporate investigations of multiple navigational tasks while quantifying MRI-derived measures of hippocampus structural integrity.

### Conclusion

We focused on navigation and spatial cognition in children with ASD. This research addresses key gaps in the existing literature on spatial abilities in the ASD population. In contrast to prior studies that typically focused on single wayfinding skills like route-learning (egocentric) or survey-learning (allocentric), our investigation encompassed multiple spatial navigation tasks included in SPACE. While children with ASD performed just as well as TD children, the presence of worse self-perceived navigational ability suggests that psychosociological confounds, such as a lack of self-belief, may exaggerate differences in navigation ability. Nonetheless, developmental differences in navigational ability were observed in our sample, with older age associated with better performance in Path Integration and Perspective Taking. These findings emphasise the importance of assessing both performance and self-perception in a navigation context, particularly in neurodevelopmental populations, where survey-based assessments of navigation may not reflect objective performance.

## Data Availability

All data produced are available online at https://github.com/MindSpaceLab/ASDSPACE

## Statements and declarations

The authors did not receive financial support from any organization for the submitted work. The authors have no competing interests to declare that are relevant to the content of this article.

## Acknowledgements

We would like to express our gratitude to all the participants involved in our study, for their support and patience, and for contributing their time.

