## Supplementary Material 1 for "Perceived vs. actual navigation ability: Differences between autistic and typically developing children"

**Tables**

*Table 1. Sociodemographic characteristics of participants*

|  | Group | N | *M* | *Median* | *SD* |
| --- | --- | --- | --- | --- | --- |
| Age (years) | ASD | - | 12.1 | 12.5 | 1.64 |
|  | TD | - | 12.0 | 13.0 | 1.59 |
| Tablet experience (years) | ASD | - | 1.72 | 2.0 | .54 |
|  | TD | - | 1.44 | 1.0 | .58 |
| Sleep per day (hours) | ASD | - | 8.5 | 9.0 | 1.25 |
|  | TD | - | 8.12 | 8.0 | 1.39 |
| Exercise per week (hours) | ASD | - | 4.58 | 3.0 | 5.57 |
|  | TD | - | 9.16 | 7.0 | 10.2 |
| Walked per week (hours) | ASD | - | 6.26 | 5.0 | 6.18 |
|  | TD | - | 13.8 | 6.5 | 20.3 |
| Handedness (% right) | ASD | 96.0 | - | - | - |
|  | TD | 88.0 | - | - | - |
| Experienced with video games (%) | ASD | 100.0 | - | - | - |
|  | TD | 84.0 | - | - | - |

*Note. ASD, Autism Spectral Disorder; TD, Typically Developed*

*Table 2. Descriptive statistics for SPACE performance in TD and ASD groups*

|  | TD | | | ASD | | |
| --- | --- | --- | --- | --- | --- | --- |
|  | *M* | *Md* | *SD* | *M* | *Md* | *SD* |
| PI Distance Error | 125.41 | 126.71 | 56.05 | 127.09 | 114.77 | 44.06 |
| PI Angle Error | 37.85 | 37.52 | 17.94 | 34.86 | 35.16 | 11.62 |
| EP Angle Error | 63.51 | 65.42 | 21.76 | 60.54 | 60.77 | 16.93 |
| Mapping Accuracy | .46 | .40 | .29 | .59 | .59 | .31 |
| PT Angle Error | 47.22 | 47.40 | 21.82 | 50.84 | 58.34 | 22.38 |

*Note. TD, Typically-Developed Children; ASD, Autism Spectrum Disorder Children; PI, Path Integration; EP, Egocentric Pointing; PT, Perspective Taking.* Path Integration Distance Error in game units; Path Integration Angle Error, Egocentric Pointing Angle Error, and Perspective Taking Angle Error is in degrees; Mapping Accuracy is in r2.

*Table 3. Descriptive statistics for SBSOD scale items in TD and ASD groups*

| SBSOD item | TD | | | ASD | | |
| --- | --- | --- | --- | --- | --- | --- |
|  | *M* | *Md* | *SD* | *M* | *Md* | *SD* |
| Total score | 4.36 | 4.27 | .61 | 3.67 | 3.67 | .69 |
| *Q1* “*I am very good at giving directions”* | 5.32 | 5.00 | .75 | 3.72 | 4.00 | 1.43 |
| *Q2* *“I have a poor memory for where I left things”* | 3.64 | 4.00 | 1.66 | 3.16 | 3.00 | 1.97 |
| *Q3 “I am very good at judging distances”* | 4.32 | 5.00 | 1.75 | 4.20 | 4.00 | 1.61 |
| *Q4 “My “sense of direction” is very good”* | 5.40 | 6.00 | .96 | 4.60 | 5.00 | 1.71 |
| *Q5 “I tend to think of my environment in terms of cardinal directions (N, S, E, W)”* | 4.04 | 4.00 | 1.97 | 2.16 | 1.00 | 1.62 |
| *Q6 “I very easily get lost in a new city”* | 3.48 | 3.00 | 1.90 | 2.72 | 2.00 | 1.72 |
| *Q7 “I enjoy reading maps”* | 3.52 | 3.00 | 1.96 | 2.76 | 2.00 | 1.96 |
| *Q8 “I have trouble understanding directions”* | 4.60 | 5.00 | 1.55 | 3.60 | 4.00 | 1.83 |
| *Q9 “I am very good at reading maps”* | 3.84 | 4.00 | 1.28 | 3.56 | 4.00 | 1.96 |
| *Q10 “I don’t remember routes very well while riding as a passenger in a car”* | 5.44 | 6.00 | 1.71 | 4.56 | 5.00 | 2.20 |
| *Q11 “I don’t enjoy giving directions”* | 4.12 | 4.00 | 1.81 | 4.44 | 5.00 | 1.80 |
| *Q12 “It’s not important to me to know where I am”* | 5.44 | 6.00 | 1.61 | 5.76 | 6.00 | 1.51 |
| *Q13“I usually let someone else do the navigational planning for long trips”* | 2.64 | 2.00 | 1.25 | 2.00 | 2.00 | 1.38 |
| *Q14 “I can usually remember a new route after I have travelled it only once”* | 4.56 | 5.00 | 1.80 | 3.64 | 4.00 | 2.10 |
| *Q15 “I don’t have a very good “mental map” of my environment”* | 5.00 | 5.00 | 1.50 | 4.16 | 4.00 | 1.84 |

**Figure 1 top**

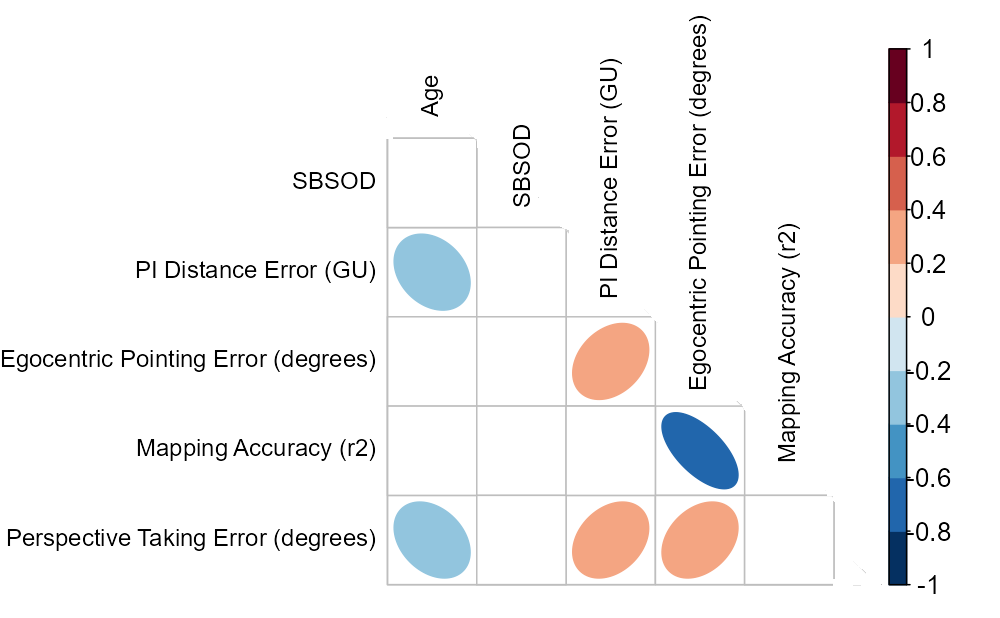

***Figure 1.*** Pearson correlations of Age, SBSOD score, and performance measures of the SPACE task. Ellipsis colour and shape represent the direction and strength of correlation. Only significant correlations are presented (*p* < .05).
